# What if risk factors influenced the variability of health outcomes as well as the mean? Evidence and implications

**DOI:** 10.1101/2021.03.30.21254645

**Authors:** David Bann, Tim J Cole

## Abstract

Risk factors may affect the variability as well as the mean of health outcomes. Understanding this can aid aetiological understanding and public health translation, in that interventions which shift the outcome mean and reduce variability are preferable to those which affect only the mean. However, few statistical tools routinely test for differences in variability. We used GAMLSS (Generalised Additive Models for Location, Scale and Shape) to investigate how multiple risk factors (sex, childhood social class and midlife physical inactivity) related to differences in health outcome mean and variability. The 1970 British birth cohort study was used, with body mass index (BMI; N = 6,025) and mental wellbeing (Warwick-Edinburgh Mental Wellbeing Scale; N = 7,128) as outcomes. For BMI, males had a 2% higher mean than females yet 28% lower variability. Lower social class and physical inactivity were associated with higher mean and higher variability (6% and 13% respectively). For mental wellbeing, gender was not associated with the mean while males had 4% lower variability. Lower social class and physical inactivity were associated with lower mean yet higher variability (−7% and 11% respectively). This provides empirical support for the notion that risk factors can reduce or increase variability in health outcomes. Such findings may be explained by heterogeneity in the causal effect of each exposure, by the influence of other (typically unmeasured) variables, and/or by measurement error. This underutilised approach to the analysis of continuously distributed outcomes may have broader utility in epidemiological, medical, and psychological sciences.

## Introduction

What is health? Contrary to simplistic notions of it being defined as the absence of disease, it is now increasingly understood that most outcomes of public health significance are continuous in nature.^1^ This applies to both physical and mental health outcomes.^2 3^ The use of binary endpoints, while having utility in clinical applications, should not hinder investigation of the influences of health outcomes which are ultimately continuous. Analysing the determinants of health using continuous rather than binary outcomes is beneficial both practically (with more statistical power and less information loss) and substantively (greater aetiological understanding).

Studies into the effect on continuous outcomes of exposures, be they risk factors in observational studies or interventions in randomised trials,^3^ typically focus on mean differences in the outcome, using linear regression. However linear regression assumes homoscedasticity, i.e. that the variability of the outcome is unrelated to the exposure, and often this is not the case. It is possible to extend the regression analysis to model the variability as well as the mean, and this has benefits in terms of not only the model’s fit but also its interpretation. If for example the intervention in an intervention trial can be shown to reduce variability in the outcome, this could reasonably be viewed as evidence of intervention success^4^ that is independent of the intervention’s effect on the mean. Treatment for refractive vision errors—glasses, contact lenses, and/or corrective surgery—seeks to improve vision by shifting individuals towards a specified standard (eg, 20/20 vision).^5^ Successful treatments alter the mean refraction, but they are even more successful if they also reduce the substantial variability in refraction arising from the mix of short- and long-sighted individuals.

Similarly, obesity interventions aim to reduce body mass index (BMI) and shift treated individuals from overweight (25-30 kg/m^2^), obese (>30 kg/m^2^) or severely obese (>45 kg/m^2^) to the normal range (20-25 kg/m^2^). However here the effect of the intervention on variability is often to increase it. Even if not formally tested, visual comparisons of outcome distributions of some influential trials suggest that weight loss interventions increase rather than reduce BMI variability,^6^ presumably since they are effective in some but not all participants.

Understanding if and how risk factors influence variability in health outcomes has aetiological significance, consistent with the goal of epidemiological science to understand the *distribution* of health.^7^ Risk factors could feasibly affect outcome variability yet not affect the mean—for example, one study found that breastfeeding was not related to mean childhood BMI, yet was related to lower childhood BMI variability.^8^ Identifying associations between risk factors and outcome variability may also be useful to identify the absence or presence of heterogeneity in susceptibility to interventions or risk factors. Indeed, the finding that substantial increases in mean BMI in recent decades have been matched by increases in BMI variability indicates that there may be differential susceptibility to the obesogenic environment.^9 10^

Recent studies in biological,^11 12^ environmental^13^ and economic science^14 15 16^ have begun to examine how risk factors relate to the distribution of the outcome of interest. However, there have been few epidemiological applications of this approach to date;^17^ and fewer still that provide explanations for such findings, which are essential if such methods are to have utility. Indeed, one recent study which investigated the association between mental health symptoms and lower income explicitly avoided interpretation of its findings on variability, focusing instead on issues relating to the application of such methods.^15^

Regression methods that allow variability to be modelled are uncommon. One particular method, Generalised Additive Models for Location, Scale and Shape (GAMLSS)^18^ has become the standard for constructing growth reference centiles,^19^ where the aim is to model the outcome’s distribution as a function of age. It defines the distribution in terms of distribution moments, i.e. the mean, variance, and optionally skewness and kurtosis. This allows for factors influencing the higher moments to be identified in just the same way as for the mean, and it provides a simple and elegant interface for modelling variability in epidemiology.

Another arguably underutilised^20^ and related statistical approach to investigating risk factors for continuous outcomes is quantile regression. Recent epidemiological studies using this method have found that risk factors for higher BMI—particularly lower social class and physical inactivity—have sizably larger effect sizes at higher BMI centiles.^21 22^ This has potentially important policy implications—risk factors which have larger effects amongst those at highest health risk are likely to have a more favourable effect on population health than alternatives which do not.^21^ However, the reason for this phenomenon is not yet understood—it may be logically consistent with results of GAMLSS analyses in which risk factors influence outcome means, variability and/or skewness.

Our primary aim is explore factors affecting outcome variability in an epidemiological context. We investigate whether and how several established risk factors—sex, socioeconomic circumstances, and physical inactivity^23^—relate to differences in outcome mean and variability. We use two different continuous outcomes as examples, an indicator of adiposity (body mass index, BMI) and mental wellbeing. A second aim is to investigate whether these risk factors are additionally related to the skewness of the outcome distribution. Finally, we investigate the same risk factor and outcome associations using quantile regression models.

## Methods

### Study sample

The 1970 British birth cohort study (1970c) consists of all 17,196 babies born in Britain during one week of March 1970, with 10 subsequent waves of follow-up from childhood to midlife.^24^ At the most recent wave (46 years), 12,368 eligible participants (those alive and not lost to follow-up) were invited to be interviewed at home by trained research staff—8,581 participants provided at least some data in this wave. At all waves, informed consent was provided and ethical approval granted.

### Health outcomes

We selected two outcomes in midlife which capture different dimensions of health and are continuously distributed: adiposity (BMI), and mental wellbeing (Warwick-Edinburgh Mental Wellbeing Scale (WEMWBS)). BMI was measured at 46 years, and wellbeing at 42 years.^25^ WEMWBS consists of 14 positively worded items—such as “I’ve been feeling optimistic about the future” and “…feeling cheerful”—measured on a five-point Likert scale, which are summed to give a total well-being score ranging from 14 to 70 (highest well-being).^26^

### Risk factors

We chose three risk factors across different domains—each of them likely to independently influence health outcomes.^23^ They were coded as binary variables to simplify comparison of descriptive and GAMLSS results: sex (female/male), socioeconomic position (social class at birth; coded as non-manual/manual), and a behavioural risk factor (reported physical activity at 42 years; reported days in which the participant took part in exercise for 30 mins or more in a typical week ‘working hard enough to raise your heart rate and break into a sweat’, coded as active (≥1 days)/inactive (0 days)). We examined if the binary split of risk factors influenced the inferences drawn—additional analyses were conducted with them coded instead as categorical variables (social class in 6 categories and physical inactivity from 0-7 days).

### Analytical strategy

To visually inspect the outcome distributions and their differences across risk factor groups, we first plotted separate kernel density estimates alongside relevant descriptive statistics (mean, standard deviation, and coefficient of variation (SD/mean)). We then used GAMLSS^18^ separately with each outcome, to formally investigate whether risk factors were associated with 1) differences in mean outcome and 2) differences in outcome variability.

GAMLSS requires the underlying distributional family to be specified—in primary analyses we used the normal distribution (NO family), where the mean and standard deviation are modelled. By convention, variability is modelled on the log scale, where differences are fractional and can be multiplied by 100 and interpreted as percentage differences in variability.^27^ Mean differences were also expressed as percentage differences to aid comparability across outcomes and model estimates.

We also used an alternative distribution family: the Box-Cox Cole and Green (BCCG family).^28^ This enabled us to examine the median (rather than the mean) and generalised coefficient of variation (rather than the SD), and in addition whether risk factors were associated with the skewness of the outcome distribution. Skewness here is quantified in terms of the Box-Cox power; the power transformation required to transform the outcome distribution to normality, where 1 indicates normal, 0 log-normal and −1 inverse normal. A smaller Box-Cox power corresponds to more right skewness.

To aid comparison of descriptive statistics and model estimation results, we first conducted analyses adjusting for each risk factor alone. We then adjusted for the risk factors jointly.

Separately we fitted conditional quantile regression models to estimate risk factor and BMI associations at the lower, middle and upper quartiles of the outcome distribution, i.e. the 25^th^, 50^th^ and 75^th^ centiles.

All analyses were conducted using Stata v16 (StataCorp, Texas), and the *gamlss* package version 5.2-0 in Rstudio 1.4.^29^

## Results

6,025 participants had valid data for BMI and all risk factors, and 7,128 for WEMWEBS. Mean BMI was 28.4 (SD = 5.5), and mean WEMWEBS 49.2 (8.3). Higher BMI was weakly associated with lower wellbeing (r = −0.08). BMI was moderately right-skewed (Figure 1) and WEMWEBS left-skewed (Figure 2). Visual and descriptive comparisons of the BMI and wellbeing distributions by risk factor (Figures 1 and 2) suggest that differences in the outcome mean and variability are not always in the same direction.

**Figure 1.**
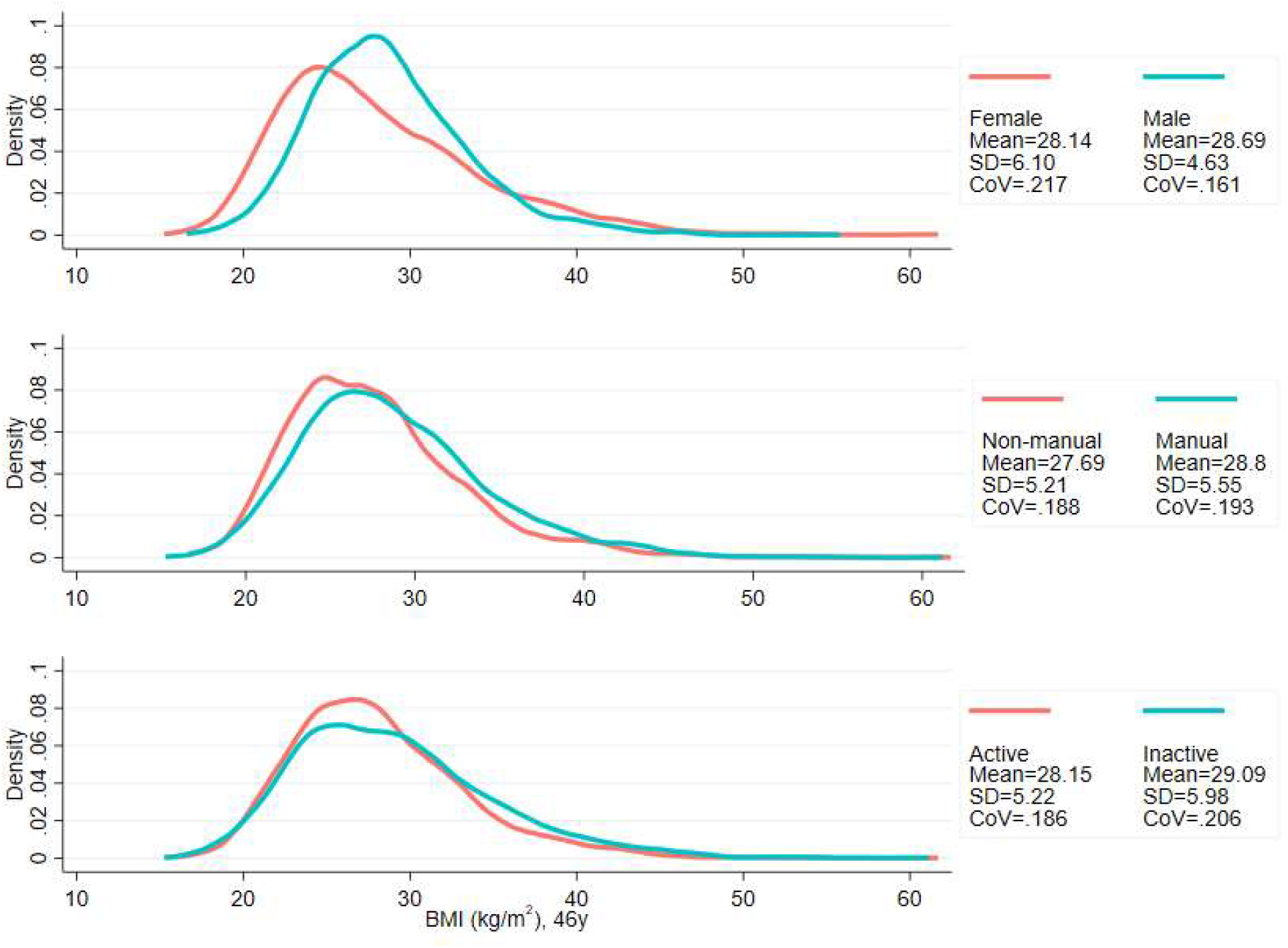
Kernel density plots for body mass index, stratified by risk factor group. Note: CoV = coefficient of variation (SD/mean).

**Figure 2.**
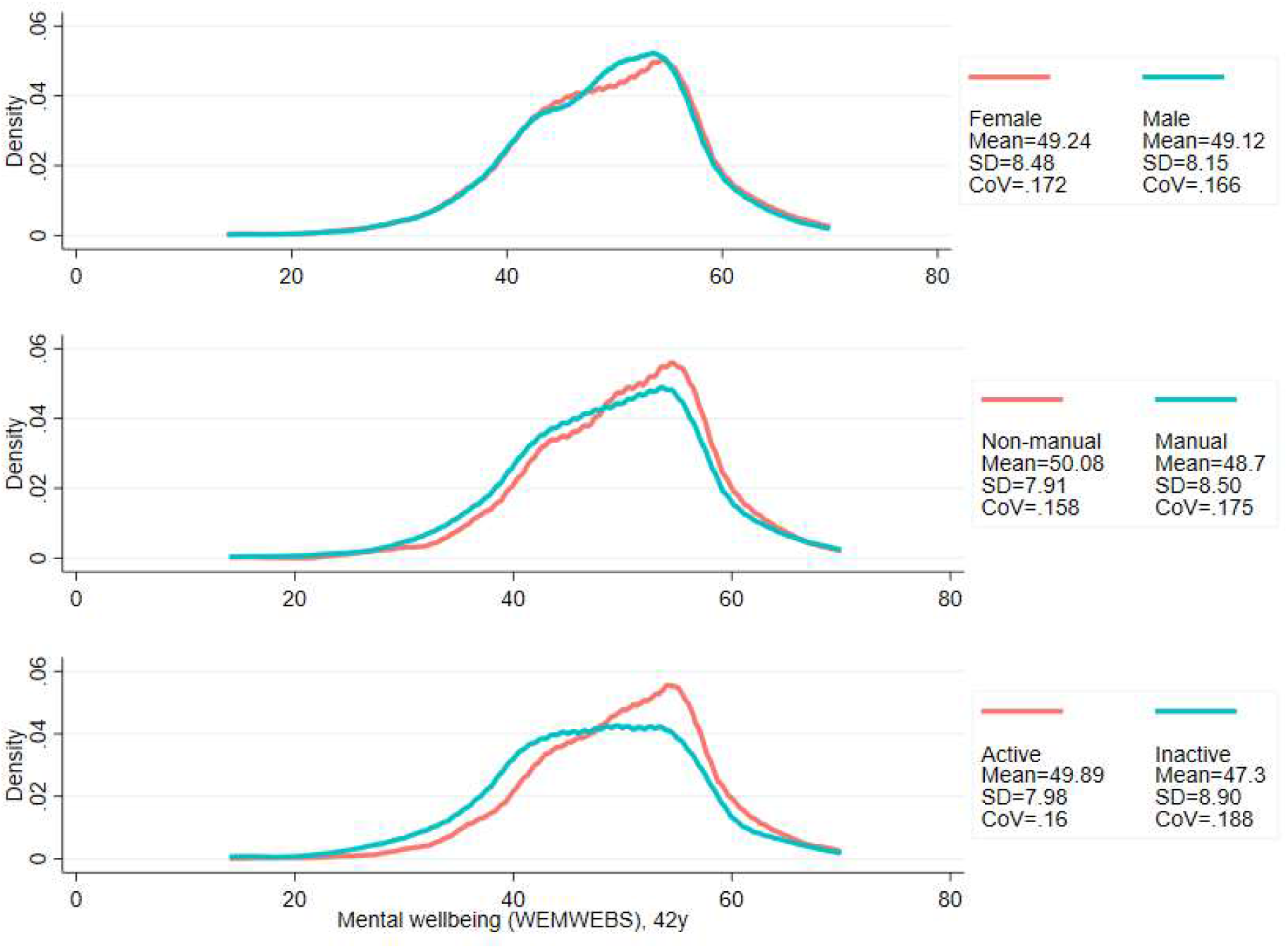
Kernel density plots for mental wellbeing, stratified by risk factor group. Note: COV = coefficient of variation (SD/mean).

GAMLSS results for the binary risk factors are shown in Tables 1 and 2, with the results using the extra risk factor categories in Supplementary Tables 1 and 2. Associations were similar in the unadjusted and mutually adjusted analyses, so the former are described below.

**Table 1.** **Risk factors in relation to body mass index: differences in mean, variability and skewness estimated by GAMLSS (n = 6,025)**

| <u>Risk factor</u> | <u>%</u> | <u>NO distribution family</u> |  | <u>BCCG distribution family</u> |  |  |
| --- | --- | --- | --- | --- | --- | --- |
|  |  | Mean | SD | Median | CoV | Skewness* |
| Female (ref) | 52.4% | 28.1 | 6.10 | 26.9 | .217 | -0.86 |
| Male | 47.6% | 28.7 | 4.63 | 28.2 | .161 | -0.38 |
| Unadjusted difference, % (SE) |  | 1.9 (0.49) | -27.6 (1.82) | 4.1 (0.49) | -23.0 (1.87) | 0.48 (0.12) |
| Adjusted<sup>#</sup> difference, % (SE) |  | 2.2 (0.48) | -27.4 (1.83) | 4.5 (0.49) | -22.6 (1.87) | 0.54 (0.12) |
| Non-manual (ref) | 36.3% | 27.7 | 5.21 | 27 | .188 | -0.85 |
| Manual social class | 63.7% | 28.8 | 5.55 | 28 | .193 | -0.47 |
| Unadjusted difference, % (SE) |  | 3.9 (0.51) | 6.2 (1.89) | 4.3 (0.51) | 6.1 (1.95) | 0.38 (0.12) |
| Adjusted<sup>#</sup> difference, % (SE) |  | 3.8 (0.48) | 5.6 (1.90) | 4.2 (0.49) | 5.6 (1.95) | 0.39 (0.12) |
| Physically active (ref) | 73.0% | 28.1 | 5.22 | 27.4 | .186 | -0.61 |
| Inactive | 27.0% | 29.1 | 5.98 | 28.3 | .206 | -0.52 |
| Unadjusted difference, % (SE) |  | 3.3 (0.58) | 13.5 (2.05) | 2.9 (0.60) | 10.5 (2.1) | 0.09 (0.13) |
| Adjusted<sup>#</sup> difference, % (SE) |  | 3.3 (0.56) | 12.1 (2.06) | 3.1 (0.57) | 9.4 (2.1) | 0.12 (0.13) |

<sup>#</sup>Estimates mutually adjusted for sex, social class and physical inactivity.
\*Skewness is estimated as the Box-Cox power (that is, the power required to transform the outcome to a normally distribution); differences are the absolute difference in Box-Cox power in each subgroup estimated by GAMLSS. GAMLSS estimates multiple distribution moments simultaneously; thus, differences may not exactly correspond to descriptive comparisons reported above.
NO: normal distribution family; BCCG: Box-Cox Cole and Green distribution family; SD: standard deviation; CoV: coefficient of variation (SD/mean); GAMLSS: Generalized Additive Models for Location, Scale and Shape.

**Table 2.** **Risk factors in relation to mental wellbeing (WEMWEBS): differences in mean, variability and skewness estimated by GAMLSS (n = 7,128)**

| <u>Risk factor</u> | <u>%</u> | <u>NO distribution family</u> |  | <u>BCCG distribution family</u> |  |  |
| --- | --- | --- | --- | --- | --- | --- |
|  |  | <u>Mean</u> | <u>SD</u> | <u>Median</u> | <u>COV</u> | <u>Skewness*</u> |
| Female (ref) | 52.8% | 49.2 | 8.48 | 50 | 0.172 | 1.67 |
| Male | 47.2% | 49.1 | 8.15 | 50 | 0.166 | 1.70 |
| Unadjusted difference, % (SE) |  | -0.2 (0.4) | -4.0 (1.7) | -0.3 (0.4) | -3.6 (1.9) | 0.03 (0.11) |
| Adjusted<sup>#</sup> difference, % (SE) |  | -0.6 (0.4) | -3.6 (1.7) | -0.7 (0.4) | -2.7 (1.9) | 0.00 (0.11) |
| Non-manual (ref) | 34.8% | 50.1 | 7.91 | 51 | 0.158 | 1.80 |
| Manual social class | 65.2% | 48.7 | 8.50 | 49 | 0.175 | 1.60 |
| Unadjusted difference, % (SE) |  | -2.8 (0.4) | 7.3 (1.8) | -2.9 (0.4) | 10.9 (2.0) | -0.20 (0.12) |
| Adjusted<sup>#</sup> difference, % (SE) |  | -2.5 (0.4) | 6.1 (1.8) | -2.7 (0.4) | 9.8 (2.0) | -0.24 (0.12) |
| Physically active (ref) | 72.4% | 49.9 | 7.98 | 51 | 0.160 | 1.68 |
| Inactive | 27.6% | 47.3 | 8.90 | 48 | 0.188 | 1.54 |
| Unadjusted difference, % (SE) |  | -5.3 (0.48) | 10.9 (1.9) | -5.2 (0.5) | 16.2 (2.1) | -0.12 (0.12) |
| Adjusted<sup>#</sup> difference, % (SE) |  | -5.3 (0.48) | 9.8 (1.9) | -5.1 (0.5) | 15.1 (2.1) | -0.10 (0.12) |

<sup>#</sup>Estimates mutually adjusted for sex, social class and physical inactivity.
\*Skewness is estimated as the Box-Cox power (that is, the power required to transform the outcome to a normally distribution); differences are the absolute difference in Box-Cox power in each subgroup estimated by GAMLSS. GAMLSS estimates multiple distribution moments simultaneously; thus, differences may not exactly correspond to descriptive comparisons reported above.
NO: normal distribution family; BCCG: Box-Cox Cole and Green distribution family; SD: standard deviation; CoV: coefficient of variation (SD/mean); GAMLSS: Generalized Additive Models for Location, Scale and Shape.

### Body mass index

Males had higher mean BMI yet lower variability than females—see Figure 1 and Table 1. The SD for BMI was lower in males (4.63) than females (6.10) i.e., a 27.6% difference (difference in log(SD) *100). This matches the estimate obtained from GAMLSS—males had 27.6% (SE: 1.8%) lower variability than females (Table 1).

In contrast, lower social class and physical inactivity were both associated with higher mean BMI and higher BMI variability (Figure 1 and Table 1). Those from lower social class households had 3.9% (0.5%) higher BMI than those from non-manual classes, and 6.2% (1.9%) higher variability.

Physically inactive participants had 3.3% (0.6%) higher mean BMI and 13.5% (2.0%) higher variability.

The GAMLSS results were similar with the BCCG distribution family rather than the NO family (Table 1). That is, risk factors associated with higher mean BMI and higher SD were also associated with higher median BMI and higher COV. Male sex and lower social class were both associated with less skewness of the BMI distribution; the Box-Cox power was 0.5 (0.1) higher in males and 0.4 (0.1) higher for manual social class. Physical activity was not associated with outcome skewness.

### Mental wellbeing – Warwick-Edinburgh Mental Wellbeing Scale

There was little evidence of sex differences in mean wellbeing, while males had marginally less variability than females—4.0% (1.7%) lower. Lower social class and physical inactivity were both associated with lower mean yet higher variability (Figure 1 and Table 2). Those from lower social class households had a 2.8% (0.4%) lower mean yet 7.3% (1.8%) higher variability. Physically inactive participants had 5.3% (0.48) lower mean yet 10.9% (1.9%) higher variability. These findings were similar in mutually adjusted analyses (Table 2).

The results were similar with the BCCG distribution family (Table 2). There was evidence suggesting that lower social class was associated with less skewness in the wellbeing distribution; sex and physical activity were not associated with outcome skewness.

### Comparison with quantile regression findings

For BMI, the associations of lower social class and physical inactivity were stronger at upper quantiles (Table 3; e.g., manual social class had 3.37 (0.35) higher BMI at the 25^th^ centile, 4.06 (0.54) at the median, and 4.82 (0.69) at the 75^th^); estimates at higher centiles were also estimated less precisely than at lower centiles (larger SE). In contrast sex differences were present at lower centiles but absent at the 75^th^. These findings corresponded with those from GAMLSS using the BCCG family, in which all BMI centiles are plotted by risk factor group (Figure 3). This comparison highlights the utility of GAMLSS—risk factor differences in the mean, variability, and skewness can each be quantified and thus visually depicted.

**Table 3.** **Risk factors in relation to body mass index (BMI) and mental wellbeing (WEMWEBS): percentage differences at multiple points of the outcome distribution estimated by quantile regression**

| Outcome | Risk factor | 25<sup>th</sup> centile | 50<sup>th</sup> centile | 75<sup>th</sup> centile |
| --- | --- | --- | --- | --- |
| BMI | Male vs female (ref) | 7.2 (0.53) | 5.1 (0.46) | 0.1 (0.61) |
|  | Manual vs Non-manual (ref) | 3.4 (0.35) | 4.1 (0.54) | 4.8 (0.69) |
|  | Inactive vs active (ref) | 1.6 (0.56) | 3.4 (0.51) | 3.8 (0.88) |
| WEMWEBS | Male vs female (ref) | -0.0 (0.67) | 0.0 (0.58) | -0.0 (0.68) |
|  | Manual vs Non-manual (ref) | -4.5 (1.05) | -2.1 (0.97) | -1.8 (0.28) |
|  | Inactive vs active (ref) | -7.1 (0.90) | -6.2 (0.75) | -3.6 (0.89) |

Note: results show the percentage difference (log-transformed x 100) in BMI or WEMWEBS (standard errors in parenthesis) at different centiles of the outcome distribution; estimates are mutually adjusted.

**Figure 3.**
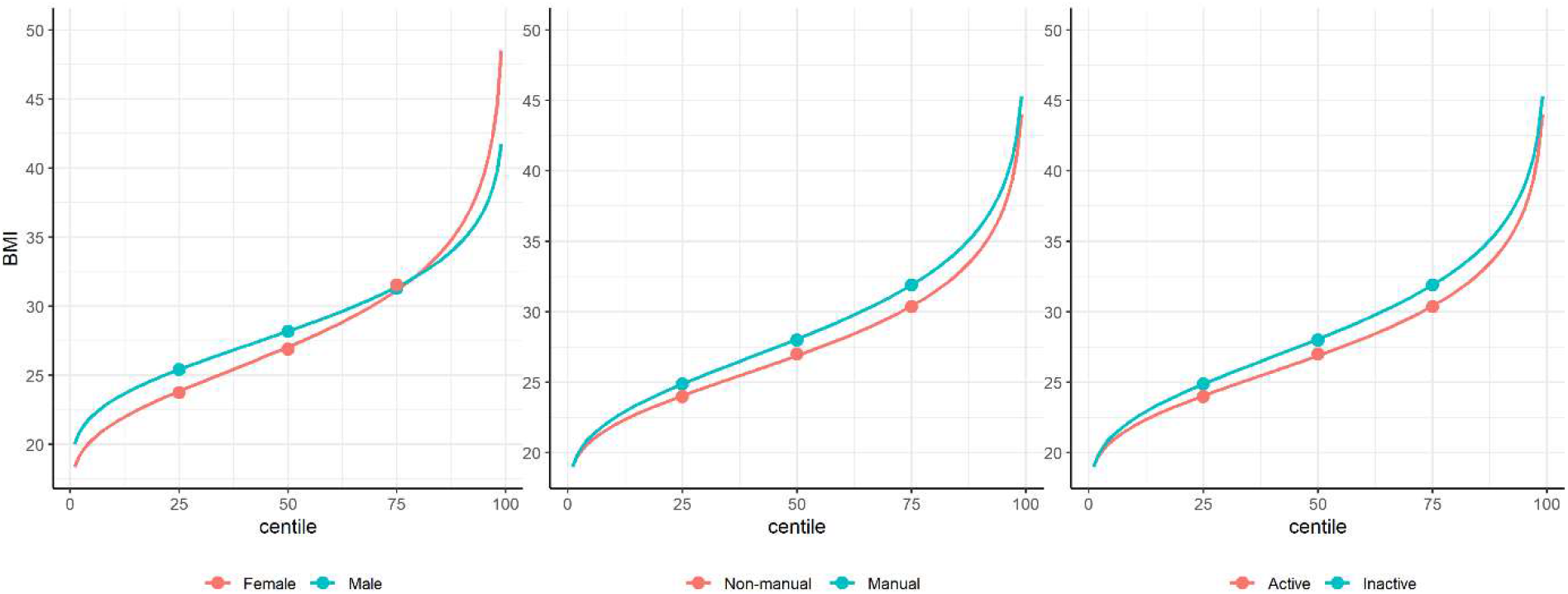
Plots of body mass index (BMI) centile by risk factor group. Plotted lines are calculated using GAMLSS estimation results of the entire outcome distribution; points at the 25^th^, 50^th^, and 75^th^ centiles are estimated using quantile regression models.

For WEMWEBS, the associations of lower social class and physical inactivity were also stronger at lower quantiles (Table 3), yet had larger SE. Sex was not associated with WEMWEBS at any centile. These findings corresponded with those from GAMLSS (Figure 4).

**Figure 4.**
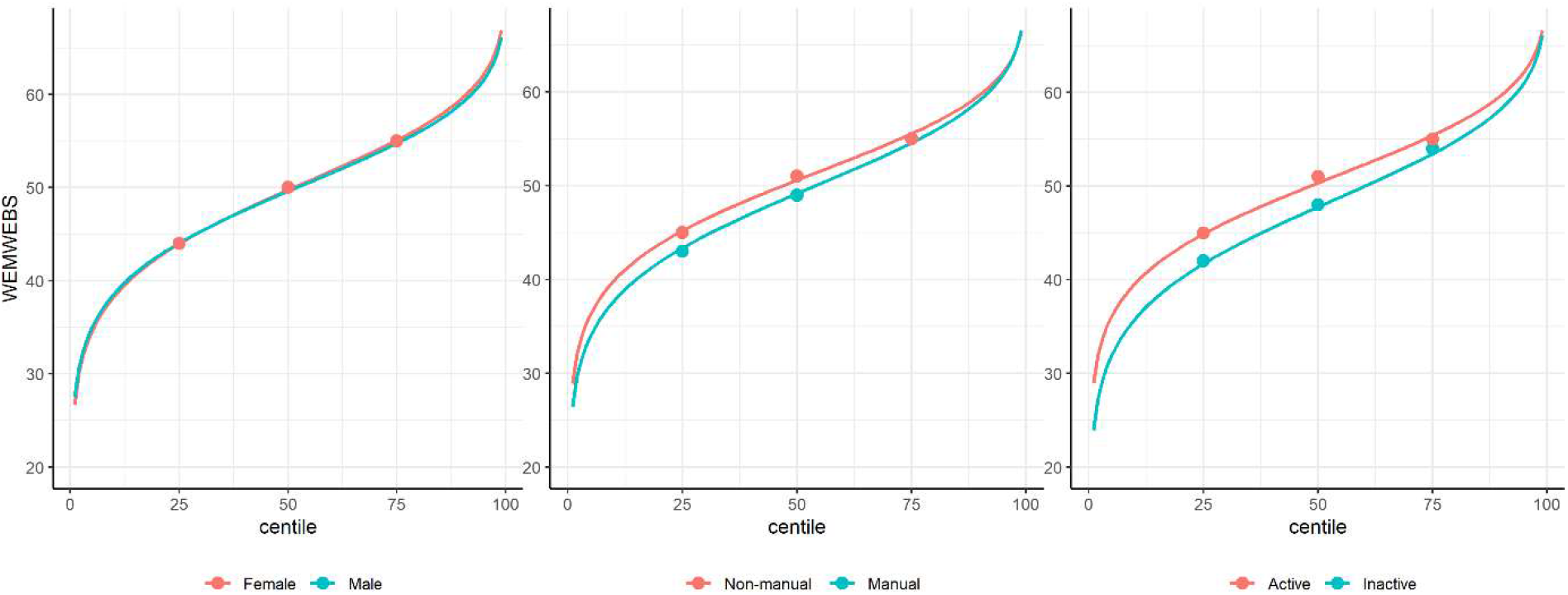
Plots of mental wellbeing (Warwick-Edinburgh Mental Wellbeing Scale, WEMWEBS) centiles by risk factor group. Plotted lines are calculated using GAMLSS estimation results of the entire outcome distribution; points at the 25^th^, 50^th^, and 75^th^ centiles are estimated using quantile regression models.

## Discussion

Using an underutilised and little-known analytical approach (GAMLSS), we present empirical evidence to support the idea that risk factors can relate to sizable differences in outcome variability, in addition to differences in the outcome mean. Females had higher variability in BMI and mental wellbeing than males; lower social class and physical inactivity were each associated with higher variability in both BMI and mental wellbeing, despite having different directions of association with the mean (higher BMI yet lower mental wellbeing).

Our findings add to an emerging literature which has investigated associations between risk factors and outcome variability. Studies^11–17^ have reported that risk factors associated with higher means are also associated with higher outcome variability. For example, Beyerlein et al (2008)^17^ found that multiple risk factors for high childhood BMI (such as more frequent television viewing and greater rapid infant weight gain) were related to both higher mean BMI and greater variability in BMI. However, previous studies have not utilised multiple outcomes or nationally representative samples, and have not systematically considered explanations for such findings or their implications.

Our findings help to reconcile findings from GAMLSS with those using quantile regression^17 21 22^ which have reported stronger effect sizes for BMI risk factors at higher BMI centiles. This finding is both consistent with and helps explain the GAMLSS findings. For instance, lower social class and physical inactivity are related to higher mean BMI, higher BMI variability, yet less BMI skewness; the net result is higher effect estimates at upper centiles which are less precisely estimated, as seen in quantile regression. While both analytical approaches have merit, GAMLSS has a number of attractive features for use in aetiological research: it enables each distribution moment to be separately investigated, and uses predetermined distribution families which enable computation of sparsely distributed variables.

Why are risk factors associated with differences in outcome variability? There are multiple possible explanations. First, it may represent the influence of unmeasured variables which influence the outcome—particularly at upper values—leading to both higher means and variability. Such additional variables could also be unmeasured effect modifiers which increases the strength of the risk factor effect at higher outcome centiles. Unmeasured factors such as genetic propensity to weight gain may for example modify the effect on weight gain of exposure to adverse socioeconomic circumstances.^30^ Other environmental factors could operate similarly—such that the association between lower social class and higher BMI is weaker amongst those living in a local environment which is less ‘obesogenic’ (i.e., conducive to more physical activity and lower energy intake).^31 32^ The net result of such divergent effects would be increased variability since the effects would range from zero to the upper bound of the effect. This explanation may also apply to mental wellbeing, given evidence for the myriad environmental^33 25^ and genetic determinants^34 35^ which could modify the effects observed in the current study.

Alternatively, between-person differences in confounding and/or measurement error may also lead to risk factors being associated with outcome variability. For example, in the present study physical activity was measured via a single item capturing reported activity of a moderate-vigorous intensity for at least 30 minutes per day; this is an imperfect reflection of the underlying exposure which may have a causal effect (e.g., total energy expenditure (across all intensities of activity) in the case of adiposity;^36^ or time spent in specific activities conducive to wellbeing in the case of mental wellbeing^37^). The net result would be higher variability in those reporting higher physical activity levels. A related issue is the extent to which the exposure captures the same ‘dose’ across participants in a given study. The physical activity measure used here counted the number of days that bouts of activity lasted at least 30 minutes; this likely reflects substantial variability in the level of exercise actually undertaken, thus leading to greater differences in outcome variability. This could partly explain the associations of lower social class with greater outcome variability, since social class is one dimension of socioeconomic position, such that there may be substantial between-person variation in other dimensions (eg, parental education, income and/or wealth^38 39^) which may each influence outcomes, leading to greater variability.

The study highlights the fact that analyses by GAMLSS and quantile regression lead to very similar results at the selected quantiles of the outcome distribution—see Figures 3 and 4. However GAMLSS, by analysing the whole distribution, can in some cases provide more efficient estimates of the quantiles. Compare for example the standard errors of the median as obtained by the BCCG family (Tables 2 and 3) and quantile regression (Table 4); for BMI the standard errors of around 0.5 are broadly similar the two ways, but for WEMWEBS the GAMLSS standard errors are appreciably smaller.

### Strengths and limitations

Strengths of this study include the analytical approach used (GAMLSS) to empirically investigate differences in outcome variability. While differences in variability can be informed by descriptive comparison (e.g., comparing SD values), GAMLSS additionally enables computation of estimates of precision and incorporates multivariable specifications (e.g., confounder or mediator adjustment; and inclusion of interaction terms). The use of the 1970 birth cohort data is an additional strength, enabling investigation of multiple risk factors and two largely orthogonal yet important continuous health outcomes. The national representation of this cohort is also advantageous—highly distorted sample selection can bias conventional epidemiological results (i.e., mean differences in outcomes),^40^ and may also bias comparisons of outcome variability.

The study also has limitations. As in all observational studies, causal inference is challenging despite the use of longitudinal data. Associations of social class at birth with outcomes for example could be explained by unmeasured confounding—this may include factors such as parental mental health. This is challenging to falsify empirically owing to a lack of such data collected before birth. In contrast, sex is randomly assigned at birth, and thus its associations with outcomes are unlikely to be confounded. However, sex differences in reporting may bias associations with mental wellbeing.

Physical activity and mental wellbeing were ascertained at broadly the same age, so that associations between the two could be explained by reverse causality; existing evidence appears to suggest bi-directionality of links between physical activity and both outcomes.^41 42^ Finally, attrition led to lower power to precisely estimate smaller effect sizes (e.g., gender differences in mental wellbeing) or confirm null effects. Such attribution could potentially bias associations—those in worse health and adverse socioeconomic circumstances are disproportionately lost to follow-up.^43 44^ The focus of principled approaches to handle missing data in epidemiology has been on the main parameter of interest—typically beta coefficients in linear regression models—and further empirical work is required to investigate the potential implications of (non-random) missingness for the variability and other moments of the outcome distribution.

### Potential implications

This study used an underutilised approach to empirically investigate associations between risk factors and outcome variability in a single cohort study. Thus, our findings require replication and extension in other datasets across other risk factors and health outcomes. Future studies should also seek to explain their findings, and where possible falsify potential explanations. Understanding how risk factors relate to and/or cause differences in outcome variability is not a standard part of epidemiological training, and it entails additional analytical and conceptual complexity. Thus, with greater application of these tools an emerging consensus on best practice should develop. In the first instance we recommend both descriptive and formal investigation, and that analysts carefully consider the use of both absolute (e.g., SD) and relative (e.g., CoV) differences in variability. Since the CoV is fractional standard deviation (eg, SD/mean or log SD), its suitability of use depends on the *a priori* anticipated relationship between the mean and variance.

In the context of randomised controlled trials, the finding of variability in treatment effects between individuals has been used to justify individualised approaches to treatment (personalised medicine). It is beyond the scope of the current article to discuss the tractability of this for complex outcomes in which treatment effects are unpredictable.^45^ Trials are designed typically to detect only mean differences in outcomes;^46^ nevertheless, additionally presenting outcome variability before and after treatment would be helpful to better appraise intervention effects.^4^ GAMLSS provides a useful framework with which to formally investigate this, even where the homoscedasticity assumption does not hold (i.e., where risk factors or treatment groups differ in their outcome variance). Where there are multiple potential efficacious interventions, further studies could meta-analyse existing trials to identify the types of intervention which additionally reduce outcome variability.

### Conclusion

We provide empirical support for the notion that risk factors or interventions can either reduce or increase variability in health outcomes. This finding is consistent with results from quantile regression analysis where a risk factor vs outcome association is stronger (or lower) at higher outcome centiles. Such findings may be explained by heterogeneity in the causal effect of each exposure, by the influence of other (typically unmeasured) variables, and/or by measurement error. This underutilised approach to the analysis of continuously distributed outcomes may have broader utility in epidemiological, medical, and psychological sciences.

## Data Availability

Data are available on the UK Data Archive

https://beta.ukdataservice.ac.uk/datacatalogue/series/series?id=200001

## Kernel density plots for BMI and WEMWEBS by risk factor group

**Supplementary Table 1.** **Risk factors in relation to body mass index (BMI): differences in mean, variability and skewness estimated by GAMLSS (n = 6,025)**

|  |  | <u>NO distribution family</u> |  | <u>BCCG distribution family</u> |  |  |
| --- | --- | --- | --- | --- | --- | --- |
| <u>Risk factor</u> | % | Mean difference, % (SE) | SD difference, % (SE) | Median difference, % (SE) | CoV difference, % (SE) | Skewness difference* |
| <u>Social class</u> |  |  |  |  |  |  |
| I professional | 6.8 | Ref | Ref | Ref | Ref | Ref |
| II intermediate | 14.9 | 4.1 (1.01) | 16.5 (4.25) | 3.2 (1) | 9.8 (4.39) | -0.2 (0.3) |
| III non-manual | 14.7 | 4.7 (1) | 12.7 (4.25) | 4.5 (0.99) | 9.0 (4.36) | 0.1 (0.29) |
| III skilled manual | 46.1 | 6.9 (0.87) | 17.1 (3.78) | 6.7 (0.87) | 12.1 (3.87) | 0.2 (0.26) |
| IV partly skilled | 12.6 | 8.5 (1.06) | 23 (4.36) | 8.7 (1.08) | 19.1 (4.44) | 0.6 (0.29) |
| V Unskilled | 5.0 | 9.3 (1.32) | 17.8 (5.44) | 9.8 (1.37) | 14.5 (5.5) | 0.7 (0.36) |
| <u>Physical activity days/week active</u> |  |  |  |  |  |  |
| ≥6 days | 13.4 | Ref | Ref | Ref | Ref | Ref |
| 5 | 9.5 | 0.5 (0.97) | 3.2 (3.88) | 0.4 (1) | 0.6 (3.94) | -0.2 (0.26) |
| 4 | 8.0 | 0.1 (1.03) | -0.2 (4.08) | 0.1 (1.04) | -2.5 (4.15) | -0.2 (0.27) |
| 3 | 15.1 | 0.7 (0.84) | -6.8 (3.43) | 0.9 (0.87) | -8.1 (3.48) | -0.2 (0.24) |
| 2 | 15.1 | 1.7 (0.85) | -3.6 (3.43) | 1.8 (0.87) | -7 (3.5) | -0.3 (0.24) |
| 1 | 11.9 | 1.2 (0.94) | 3.5 (3.68) | 1 (0.94) | -1.3 (3.73) | -0.1 (0.24) |
| 0 days | 27.0 | 4 (0.79) | 10.7 (3.06) | 4 (0.82) | 6 (3.09) | 0 (0.2) |

Estimates mutually adjusted for sex, social class and physical inactivity.
\*Skewness is estimated as the Box-Cox power (that is, the power required to transform the outcome to a normally distribution); differences are the absolute difference in Box-Cox power in each subgroup estimated by GAMLSS. GAMLSS estimates multiple distribution moments simultaneously; thus, differences may not exactly correspond to descriptive comparisons reported above.
NO: normal distribution family; BCCG: Box-Cox Cole and Green distribution family; SD: standard deviation; CoV: coefficient of variation (SD/mean); GAMLSS: Generalized Additive Models for Location, Scale and Shape.

**Supplementary Table 2.** **Risk factors in relation to mental wellbeing (WEMWEBS): differences in mean, variability and skewness estimated by GAMLSS (n = 7,128)**

| <u>Risk factor</u> | <u>%</u> | <u>NO distribution family</u> |  | <u>BCCG distribution family</u> |  |  |
| --- | --- | --- | --- | --- | --- | --- |
|  |  | Mean difference, % (SE) | SD difference, % (SE) | Median difference, % (SE) | CoV difference, % (SE) | Skewness difference* |
| Social class |  |  |  |  |  |  |
| I professional | 6.2 | Ref | Ref | Ref | Ref | Ref |
| II intermediate | 14.4 | -1.0 (0.88) | 0.3 (4.06) | -1.1 (0.89) | 2.7 (4.53) | -0.3 (0.31) |
| III non-manual | 14.3 | -1.8 (0.88) | 0.0 (4.06) | -1.9 (0.9) | 2.7 (4.54) | -0.2 (0.31) |
| III skilled manual | 46.5 | -3 (0.79) | 5.6 (3.62) | -3.2 (0.8) | 10.6 (4.04) | -0.4 (0.27) |
| IV partly skilled | 13.6 | -5.2 (0.92) | 7.2 (4.09) | -5.6 (0.95) | 15.3 (4.56) | -0.5 (0.29) |
| V Unskilled | 5.1 | -5.7 (1.16) | 3.3 (5.05) | -5.7 (1.2) | 10.7 (5.68) | -0.3 (0.39) |
| <u>Physical activity days/week active</u> |  |  |  |  |  |  |
| ≥6 days | 13.6 | Ref | Ref | Ref | Ref | Ref |
| 5 | 9.6 | 0.8 (0.82) | -7.5 (3.54) | 0.7 (0.85) | -7.4 (3.92) | 0.0 (0.24) |
| 4 | 7.9 | 3.0 (0.86) | -6.5 (3.77) | 2.5 (0.9) | -6.8 (4.09) | -0.3 (0.26) |
| 3 | 14.9 | 1.8 (0.72) | -10 (3.14) | 1.9 (0.74) | -12.8 (3.52) | 0.3 (0.22) |
| 2 | 14.6 | 0.9 (0.72) | -12 (3.16) | 0.7 (0.75) | -11.9 (3.5) | 0.0 (0.22) |
| 1 | 11.8 | -1.8 (0.79) | -5.8 (3.34) | -2 (0.82) | -2.8 (3.7) | -0.1 (0.22) |
| 0 days | 27.6 | -4.5 (0.7) | 3.2 (2.79) | -4.5 (0.72) | 8.4 (3.12) | -0.1 (0.18) |

Estimates mutually adjusted for sex, social class and physical activity.
\*Skewness estimates are the box-cox power in each subgroup (that is, the power required to transform the outcome to a normally distributed variable); differences are the absolute difference in box-cox power in each subgroup estimated by GAMLSS.
NO: normal distribution family; BCCG: Box-Cox Cole and Green distribution family; SD: standard deviation; COV: coefficient of variation (SD/mean); GAMLSS: Generalized Additive Models for Location, Scale and Shape (GAMLSS).

